# Plasma p-tau217 in Alzheimer’s disease: Lumipulse and ALZpath SIMOA head-to-head comparison

**DOI:** 10.1101/2024.05.02.24306780

**Authors:** Andrea Pilotto, Virginia Quaresima, Chiara Trasciatti, Chiara Tolassi, Diego Bertoli, Cristina Mordenti, Alice Galli, Andrea Rizzardi, Salvatore Caratozzolo, Andrea Zancanaro, José Contador, Oskar Hansson, Sebastian Palmqvist, Giovanni De Santis, Henrik Zetterberg, Kaj Blennow, Duilio Brugnoni, Marc Suárez-Calvet, Nicholas J Ashton, Alessandro Padovani

## Abstract

**Background:** Plasma phosphorylated-tau217 (p-tau217) has been shown to be one of the most accurate diagnostic markers for Alzheimer’s disease (AD). No studies have compared the clinical performance of p-tau217 as assessed by the fully automated Lumipulse and SIMOA ALZpath p-tau217.

**Aim:** To evaluate the diagnostic accuracy of Lumipulse and SIMOA plasma p-tau217 assays for AD.

**Methods:** The study included 392 participants, 162 with AD, 70 with other neurodegenerative diseases (NDD) with CSF biomarkers and 160 healthy controls. Plasma p-tau217 levels were measured using the Lumipulse and ALZpath SIMOA assays. The ability of p-tau217 assessed by both techniques to discriminate AD from NDD and controls was investigated using ROC analyses.

**Results:** Both techniques showed high internal consistency of p-tau217 with similar correlation with CSF p-tau181 levels. In head-to-head comparison, Lumipulse and SIMOA showed similar diagnostic accuracy for differentiating AD from NDD (area under the curve [AUC] 0.952, 95%CI 0.927–0.978 vs 0.955, 95%CI 0.928–0.982, respectively) and HC (AUC 0.938, 95%CI 0.910-0.966 and 0.937, 95% CI0.907-0.967 for both assays).

**Conclusions:** This study demonstrated the high precision and diagnostic accuracy of p-tau217 for the clinical diagnosis of Alzheimer’s disease using either fully automated or semi-automated techniques.

## INTRODUCTION

Cerebrospinal fluid (CSF) biomarkers are informative, sensitive and specific for the detection of Alzheimer’s disease (AD) in clinical and research settings from early stages of the disease.^1,2^ The recent development of plasma biomarkers is dramatically changing the AD scenario, as they are scalable tools to aid clinical evaluation and trial recruitment.^3,4^ Phosphorylated tau species (p-tau) stand at the forefront of emerging AD blood tests, exhibiting superior accuracy in diagnosis and specificity for the disease compared to the Aβ42/40 ratio or other suggested biomarkers.^5–9^

To date, phosphorylated tau at threonine 217 (p-tau217) appeared to be one of the most sensitive and specific AD markers compared to other phosphorylated tau species for differentiating AD from other neurodegenerative disorders.^6,10–16^

In addition, p-tau217 exhibits a unique longitudinal trajectory in preclinical AD amyloid-positive individuals, with increases over time being significantly associated with worsening cortical atrophy and declining cognitive performance.^4,6,13,17,18^

Most published studies focusing on p-tau species have used immunoassays on either the Meso Scale Discovery (MSD) or SIMOA platforms.^5,10,11,15,19^ The recent development of similar assays using chemiluminescent enzyme immunoassay (CLEIA) technology (including the fully automated Lumipulse platform) represents an attractive further step for their easier use and wider consistent applicability in clinical practice. The fully automation produces more consistent results between laboratories, and overtime in the same laboratory.

For Lumipulse p-tau217, only one preliminary study suggested a high discrimination accuracy for AD diagnosis, thought without a head-to-head comparison available to date.^20^ Despite the growing number of unpublished data available, there is an urgent need for high quality technical and clinical validation of newly developed p-tau217 markers.

The objective of the study was therefore to compare the diagnostic accuracy performance of Lumipulse vs SIMOA plasma p-tau217 in a large real-world memory clinic scenario with clinically approved CSF AD biomarkers as the reference standard.

## MATERIAL AND METHODS

### Study population

The study included participants with mild cognitive impairment (MCI) or mild dementia who underwent CSF assessment at the outpatient Neurodegenerative clinic of the Brescia University Hospital, Italy and matched healthy control subjects. A standardized full cognitive and behavioural assessment, including Mini-Mental State Examination (MMSE), Neuropsychiatric Inventory (NPI) and Clinical Dementia Rating Scale (CDR) was performed in each participant. Each patient underwent lumbar puncture in fasting condition according to the standardized protocol of the outpatient Neurodegenerative clinic. The CSF specimens were collected in 15 mL polypropylene sterile tubes, gently mixed to avoid gradient effects and sent directly to the hospital laboratory for routine assessments and Lumipulse CSF core AD markers.^21^

Patients with cognitive impairment or dementia were further classified in AD vs neurodegenerative disorders (NDD) according to the Internal cut-off value of Lumipulse Aβ42/p-tau181 ratio >11.1.^22,23^ A group of neurologically and cognitively normal individuals (healthy controls, HC) were recruited from participants’ caregivers and was included as reference group for plasma analyses.

The study was approved by the local ethics committee (NP 1471, DMA, Brescia) and was performed in conformity with the Helsinki Declaration; informed consent was obtained from each study participant or their legally authorized representative.

### Plasma collection and analysis

Blood samples were collected from each participant using 7.5 mL tubes containing K2-ethylenediaminetetraacetic acid (K2-EDTA). The tubes were gently inverted 5 to 10 times to mix the blood and then centrifuged at 2500×g for 10 minutes at room temperature (RT). Next, 0.5mL plasma aliquots were pipetted into polypropylene cryotubes and stored at ultra-low temperature freezing (ULTF) −80°C.

On the day of analysis, the plasma samples were brought to RT (21–23 °C). Following the manufacturer’s instructions, plasma samples were centrifuged at 2000g for 5 minutes. The plasma was then transferred to the instrument cuvettes for testing on Lumipulse using the Lumipulse G p-tau217 - Plasma Immunoreaction Cartridges RUO (for research use only).

The commercial ALZpath p-tau217 assay uses a proprietary monoclonal p-tau217 specific capture antibody, an N-terminal detector antibody, and a peptide calibrator.^5^ It has been validated as a fit-for-purpose assay^24^ with a limit of detection of 0.0052 to 0.0074 pg/mL, a functional lower limit of quantification of 0.06 pg/mL, and a dynamic range of 0.007 to 30 pg/mL. The spike recovery for the endogenous analyte was 80%, and intrarun and interrun precision was 0.5% to 13% and 9.2% to 15.7%, respectively. SIMOA analyses were performed on HD-X with commercially available p-tau217 ALZpath Simoa pTau-217 V2 Kits (Quanterix) at the Clinical Neurochemistry Laboratory, Sahlgrenska University Hospital, Mölndal, Sweden.^5^

### Statistical and precision analyses

The study investigated the within-lab precision of the Lumipulse plasma Immunoreaction Cartridges RUO through repeated inter-day testing schemes 3X5 and 5×5. For the 3×5 testing, three plasma aliquots from a healthy control and three plasma aliquots from an AD patient were used. Two commercial QC samples, namely the high and the low-level controls provided by the company, were tested five times a day for five days. The Lumipulse testing precision and between-day repeatability has been assessed in 15 and 25 runs, respectively. The calculation of the intermediate precision and the between-run precision were calculated following the CLSI EP15.^25^ The 15 independent plasma samples were stored at −80°C during the 5 days of the assessments. The 25 commercial QC of the p-tau217 kit were kept at −20°C as per the manufacturer’s instructions.

Normality distribution was evaluated using the Shapiro-Wilk test and Q-Q plots, outliers were defined based on single values higher/lower than 3 standard deviations compared to the mean of the group. To compare clinical and demographic characteristics as well as cognitive assessments and CSF and plasma markers between diagnostic groups (AD, HC, NDD), Kruskal-Wallis test was conducted. Post-hoc comparisons between AD, HC and NDD was performed using the Mann-Whitney U test. The comparability between the two analytical platforms was assessed using Passing-Bablok regression, while their imprecision was assessed by calculating the laboratory’s coefficient of variation (CV). The association between plasma and CSF biomarkers was determined using Pearson’s correlation coefficient within a correlation matrix. The accuracy in discriminating between AD and NDD/HC using plasma biomarkers, in terms of specificity and sensibility, was assessed using a receiver operating characteristic (ROC) approach. Area under the ROC curve (AUCs) were computed using the pROC package in R. The same statistical analyses were performed only considering AD-MCI and NDD-MCI subgroups (i.e. CDR <1). All analyses were conducted using R Statistical Software (https://www.r-project.org/). Statistical significance was defined at α=0.05, and all tests were two-tailed.

## RESULTS

### Precision and repeatability of p-tau217 Lumipulse G600II testing

Fifteen different specimens were aliquoted from plasma samples collected from one AD CSF-confirmed patient (Positive control, PC) and a healthy control subject (Negative control, NC), both tested as independent samples to perform the between-day repeatability and calculate the testing precision. The Clinical Laboratory’s CV and the between-run CV(%) for PC and NC for p-tau217 resulted in 2.340 and 1.310 for the PC, 3.749 and 2.280 for the NC, respectively (Suppl. Tables 1,2 and 5). Likewise, commercial Quality controls (QC) materials were also used (Suppl. Table 3 and 4). The commercial samples resulted in a CV within laboratory and between run of 5.080 and 5.340, for the L1, and 3.387 and 3.490 for the L2, respectively (Suppl. Table 5).

### Clinical validation and SIMOA head-to-head comparison

The clinical study included 392 subjects, namely 232 patients and 160 controls. The clinical assessment and CSF AD markers allowed the classification of patients in 162 AD (of which 112 had MCI) and 70 other neurodegenerative disorders (of which 45 had MCI). No outliers were detected and all SIMOA and Lumipulse values were included in the final analyses. In the whole cohort and AD/NDD/HC subgroups, no correlations between age and p-tau217 levels (tested by Lumipulse and SIMOA) were detected. Clinical and demographics data and CSF core biomarkers are indicated in Table 1. P-tau217 values showed a constant, systematic and proportional error between the two detection methods as highlighted by the Passing-Bablok regression (Figure 1). The intercept is 0.067 (95%CI 0.046 – 0.084) and the slope=1.552 (95%CI 1.433 – 1.703). AD showed higher levels of plasma p-tau217 assessed with both techniques compared to both NDD and HC (Table 1 and Figure 1). The correlation analyses demonstrated a positive relationship between plasma p-tau217 analysed by Lumipulse testing and CSF p-tau181 and t-tau (respectively r=0.578, p<0.001 and r=0.431, p<0.001). A similar positive correlation was found for plasma p-tau217 tested by SIMOA (r=0.595, p<0.001 and r=0.659, p<0.001), being p-tau217 Lumipulse/SIMOA levels highly correlated (r=0.830, p<0.001).

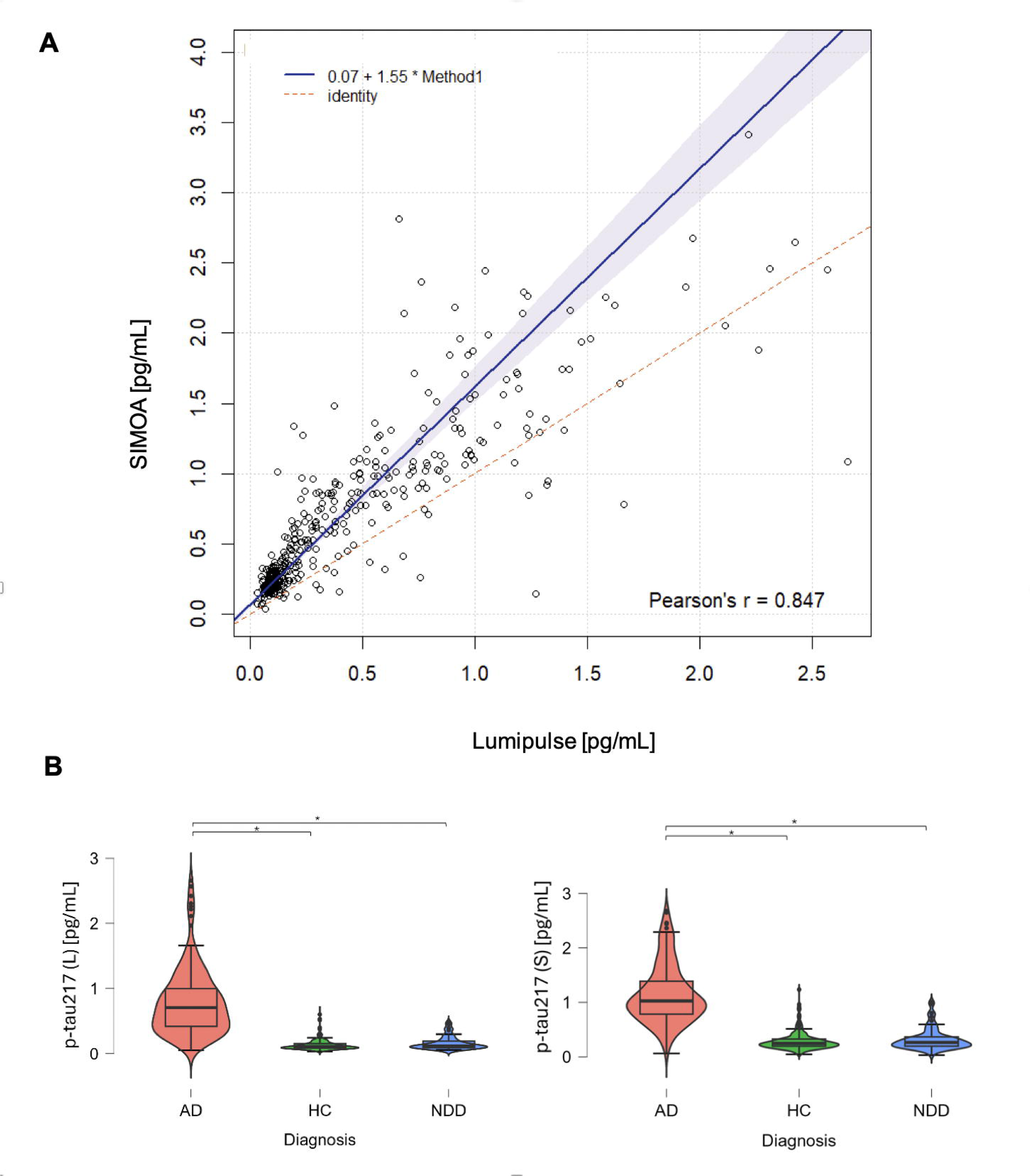

**Table 1.** Participants’ characteristics and Plasma biomarkers assessed by Lumipulse and SIMOA platforms.

|  | **HC**<br>**(N=160)** | **AD**<br>**(N=162)** | **NDD**<br>**(N=70)** | **p-value** |
| --- | --- | --- | --- | --- |
| Age | 71.016 (5.450) | 72.478 (7.287) | 69.593 (7.332) | 0.005<sup>a</sup> |
| Sex (F:M) | 101:59 | 101:57 | 28:42 | <0.001<sup>a</sup> |
| MMSE | 28.70 (1.0) | 23.083 (4.790) | 24.500 (5.651) | <0.001<sup>b, c</sup> |
| **AD CSF core biomarkers** |  |  |  |  |
| t-tau (pg/mL) | - | 696.481 (388.766) | 360.904 (257.172) | <0.001 |
| p-tau181 (pg/mL) | - | 113.943 (61.599) | 40.007 (15.040) | <0.001 |
| Aβ42 (pg/mL) | - | 479.332 (165.290) | 1060.177 (1051.273) | <0.001 |

| Plasma biomarkers |  |  |  |  |
| --- | --- | --- | --- | --- |
| Plasma p-tau217 (L) (pg/mL) | 0.181 (0.222) | 0.794 (0.511) | 0.163 (0.105) | <0.001<sup>a, b</sup> |
| Plasma p-tau217 (S) (pg/mL) | 0.353 (0.349) | 1.163 (0.565) | 0.323 (0.196) | <0.001<sup>a, b</sup> |

Data are expressed as mean (M) and standard deviation (SD). p-values showed the difference between AD CSF core biomarkers profile groups and were computed with a Mann-Whitney U test (age, MMSE, AD CSF core biomarkers), or a chi-square (sex). **Abbreviations:** AD, Alzheimer's disease; HC, healthy control; MMSE, Mini-Mental State Examination; mL, millilitres; NDD, non-Alzheimer neurodegenerative disorders; p-tau181, phosphorylated tau 181 isoform; p-tau217, phosphorylated tau 217 tested on Lumipulse (L) and SIMOA (S); Aβ42, amyloid beta 1–42; pg, picograms. <sup>a</sup> = significant comparison AD *versus* NDD; <sup>b</sup> = significant comparison AD *versus* HC; <sup>c</sup> = significant comparison NDD *versus* HC.

### Discriminant analyses for AD diagnosis

The discrimination accuracy of plasma biomarkers analysed with Lumipulse and SIMOA techniques for the diagnosis of AD with respect to both HC and NDD was separately evaluated using AUC-ROC analysis (Figure 2). Plasma p-tau217 analysed on the Lumipulse system, resulted in an AUC for AD vs NDD of 0.952 (95%CI 0.927–0.978) and 0.938 (95%CI 0.910–0.966) vs HC. Plasma p-tau217 tested on SIMOA yielded similar diagnostic accuracy, with an AUC of 0.955 (95%CI 0.928–0.982) for the discrimination of AD from NDD and 0.937 (95%CI 0.907–0.967) from HC. The calculated best cutoffs (i.e highest Youden index) for AD vs HC and AD vs NDD were 0.291 pg/mL and 0.276 pg/mL (Figure 2), respectively, for Lumipulse. The computed best cutoffs considering p-tau217 levels in SIMOA, for AD vs HC and AD vs NDD were 0.542 pg/ml and 0.518 pg/ml, respectively, (highest Youden index).

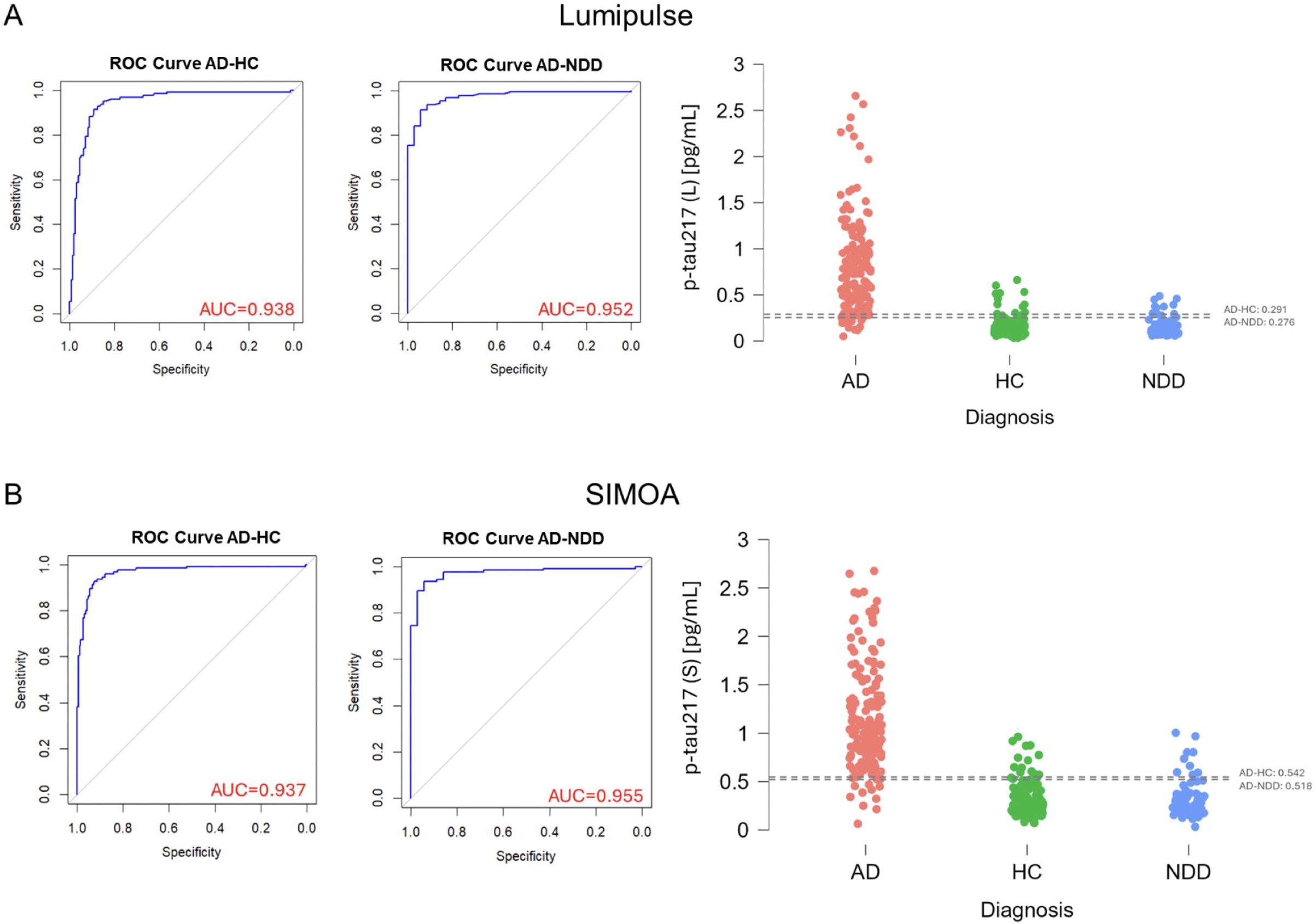

In the MCI subset, including 112 AD-MCI and 45 NDD-MCI, the AUC and the cutoffs were similar to the whole cohort (Suppl. Tables 6 and 7). Specifically, Lumipulse p-tau217 yielded an AUC of 0.946 (95%CI 0.911–0.981) for the discrimination between AD-MCI and NDD-MCI and of 0.960 (95%CI 0.936–0.985) for the differentiation with HC. SIMOA ALZpath p-tau217 exhibited a similar accuracy with AUCs of 0.934 (95%CI 0.893–0.976) vs NDD-MCI and 0.960 (95%CI 0.936–0.985) for HC (Suppl. Table 7).

## DISCUSSION

This study demonstrated the excellent clinical accuracy of plasma p-tau217 for AD detected using Lumipulse and SIMOA techniques. These suggest that both techniques are valid, solid and comparable alternatives for the assessment of plasma p-tau217 levels, potentially broadening the accessibility of this biomarker in clinical settings.

The technical validation of Lumipulse p-tau217 assessment showed a CV within laboratory around 5% for p-tau217 lower levels, otherwise below 3.5% for higher level plasma samples. These values are in line with the precision levels observed for both SIMOA and MSD techniques.^5,15^ The method comparison analysis (Passing-Bablok) showed that the two testing platforms identified different but highly related p-tau217 concentrations. Therefore, two distinct cut-offs (or conversion methods) for p-tau217 are required for Lumipulse and SIMOA techniques. This is consistent with previous data evaluating p-tau181 assays across techniques in clinical settings.^21^

When applied in a clinical setting, the p-tau217 plasma assay confirmed its high biological validity, with a high discrimination accuracy of more than 93% for AD compared to other CSF-confirmed neurodegenerative diseases and age-matched healthy controls. These results are consistent with the greater fold change of p-tau217 compared to other p-tau species, namely p-tau231 and p-tau181 recently demonstrated.^4,5,10–16^

Of note, the cutoffs resulting in the highest Youden index in the ROC analyses for discriminating AD versus NDD and controls resulted in very similar cutoff values across assays, suggesting the possible adoption of a single value for AD diagnosis, ideally to be established by multi-centre validation studies. The head-to-head comparison with ALZpath p-tau217 showed a slightly higher fold-change for Lumipulse with a comparable discrimination accuracy compared to SIMOA, even in the subcohort of MCI subjects at earlier stages of the disease.

The strong correlation between plasma p-tau217 and CSF p-tau181 standard levels further supports its utility as a non-invasive alternative for diagnosis and monitoring of Alzheimer’s disease, potentially limiting CSF analysis to a subset of subjects with borderline levels.^26^ Noteworthy, the study included subjects with different diseases and ages not accounting on *a priori* selection, thus confirming the broad applicability of such techniques in real-life settings. Nevertheless, further technical validations of the testing methods are warranted to challenge the stability of biomarkers *in vivo* in different settings, as testing immediately after −80°C storage is not always available. This is particularly important when considering the transition from research to clinical use of such assays.^21,27,28^ While our study demonstrates high concordance between the Lumipulse and SIMOA techniques, further validation efforts are warranted to confirm the biological relevance of plasma p-tau217 as a reliable biomarker for Alzheimer’s disease in different patient populations and disease stages, even using different standard-of-truth methods.

Future research should focus on addressing the remaining validation gaps by using predefined cut-offs values and optimising the clinical utility of plasma p-tau217 assays. Furthermore, longitudinal studies are needed to establish the stability over short and long-term time window and the prognostic value of plasma p-tau217 in predicting disease progression and treatment response in AD patients, even in combination with other existing plasma biomarkers. In addition, efforts should be made to standardise assay protocols and establish reference ranges for plasma p-tau217 levels to facilitate its integration into routine clinical practice for early detection and monitoring of AD continuum.

In conclusion, our study adds to the growing body of evidence supporting the utility of plasma p-tau217 as a reliable biomarker for the diagnosis of Alzheimer’s disease. The validation of Lumipulse p-tau217 highlights its potential to complement existing diagnostic approaches and improve the accuracy of AD detection in clinical practice.

## DATA AVAILABILITY

The datasets used and analyzed during the current study are available from the corresponding author on reasonable request.

## ACKNOWLEDGEMENTS

The authors wish to thank the study participants who took part in this research. We are very grateful for the assistance received from all the nurses of the Neurology department of the Spedali Civili of Brescia. We want to thank especially the laboratory technicians of the Central Clinical Laboratory for their amazing availability.

## AUTHORS’ CONTRIBUTION

Api, VQ: contributed to design the work, acquisition, analysis, interpretation of data. CTo, MP, DB, CM, AG, AR, AZ: data acquisition and analysis. CTr, VQ: data analysis and statistics. APa, DBr: have drafted the work and substantively revised it. SC, AB, KB, HZ, SG, NJA, MSC, JC, OH, SP, GDS: revised and approved the submitted version.

## SUPPLEMENTARY MATERIAL

Supplementary material include 6 tables and 3 figures.

---

**Table 1.**
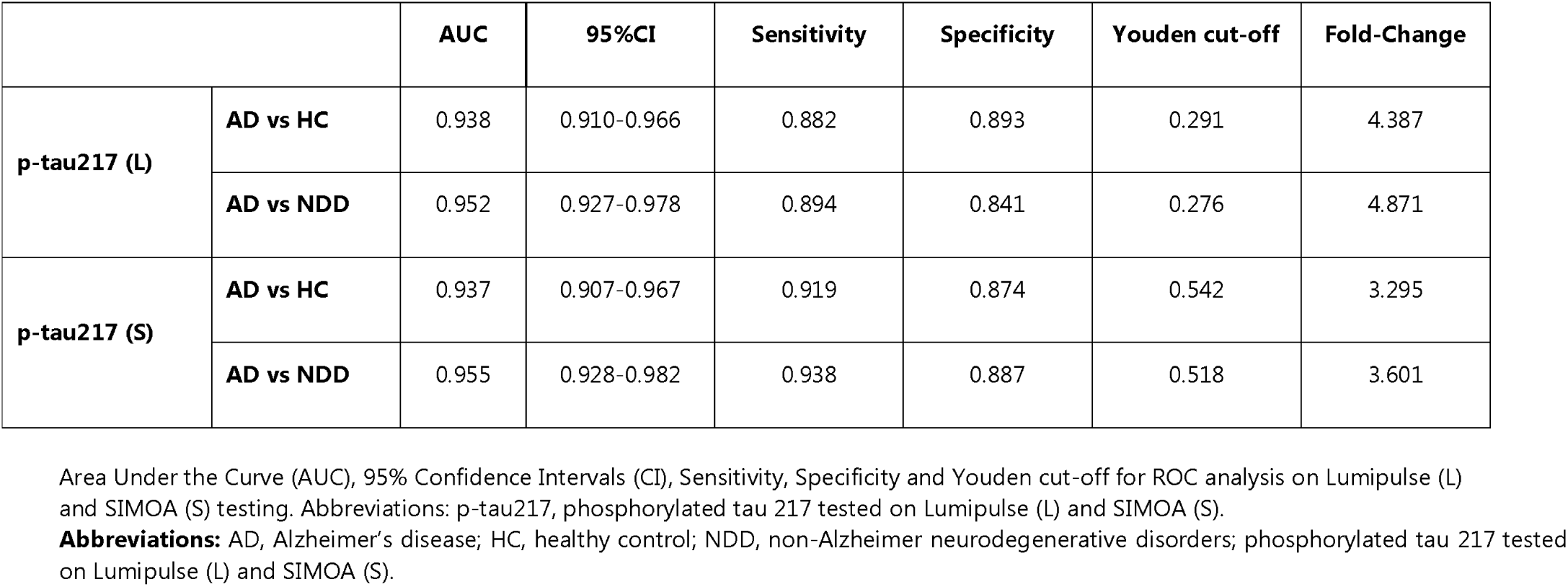
Diagnostic accuracy of Lumipulse (L) and Simoa (S) plasma p-tau217

|  |  | AUC | 95%CI | Sensitivity | Specificity | Youden cut-off | Fold-Change |
| --- | --- | --- | --- | --- | --- | --- | --- |
| p-tau217 (L) | AD vs HC | 0.938 | 0.910-0.966 | 0.882 | 0.893 | 0.291 | 4.387 |
|  | AD vs NDD | 0.952 | 0.927-0.978 | 0.894 | 0.841 | 0.276 | 4.871 |
| p-tau217 (S) | AD vs HC | 0.937 | 0.907-0.967 | 0.919 | 0.874 | 0.542 | 3.295 |
|  | AD vs NDD | 0.955 | 0.928-0.982 | 0.938 | 0.887 | 0.518 | 3.601 |

Area Under the Curve (AUC), 95% Confidence Intervals (CI), Sensitivity, Specificity and Youden cut-off for ROC analysis on Lumipulse (L) and SIMOA (S) testing. Abbreviations: p-tau217, phosphorylated tau 217 tested on Lumipulse (L) and SIMOA (S).
**Abbreviations:** AD, Alzheimer's disease; HC, healthy control; NDD, non-Alzheimer neurodegenerative disorders; phosphorylated tau 217 tested on Lumipulse (L) and SIMOA (S).

