## Supplementary tables 1-7 for "Plasma p-tau217 in Alzheimer’s disease: Lumipulse and ALZpath SIMOA head-to-head comparison"

**Supplementary Table 1.** **Repeatability test Negative control, NC (3 replicates X 5 testing days)**

| p-tau217 LUMIPULSE | NC-1 | NC-2 | NC-3 | MEAN | SD | CV(%) |
| --- | --- | --- | --- | --- | --- | --- |
| day 1 (pg/mL) | 0.170 | 0.176 | 0.181 | 0.176 | 0.006 | 3.135 |
| day 2 (pg/mL) | 0.189 | 0.186 | 0.186 | 0.187 | 0.002 | 0.926 |
| day 3 (pg/mL) | 0.189 | 0.194 | 0.188 | 0.190 | 0.003 | 1.689 |
| day 4 (pg/mL) | 0.184 | 0.194 | 0.183 | 0.187 | 0.006 | 3.253 |
| day 5 (pg/mL) | 0.177 | 0.181 | 0.182 | 0.180 | 0.003 | 1.470 |

repeatability test of the Lumipulse G600II using 3 aliquots of a healthy controls (Negative control, NC) tested in 5 different days. p-tau217, phosphorylated tau 217; SD, Standard Deviation; CV, Coefficient of Variation; 95%CI

**Supplementary Table 2. Repeatability test Positive control, PC (3 replicates X 5 testing days)**

| p-tau217 LUMIPULSE | PC-1 | PC-2 | PC-3 | MEAN | SD | CV(%) |
| --- | --- | --- | --- | --- | --- | --- |
| day 1 (pg/mL) | 0.480 | 0.494 | 0.497 | 0.490 | 0.009 | 1.851 |
| day 2 (pg/mL) | 0.479 | 0.479 | 0.478 | 0.479 | 0.001 | 0.121 |
| day 3 (pg/mL) | 0.470 | 0.479 | 0.473 | 0.474 | 0.005 | 0.967 |
| day 4 (pg/mL) | 0.484 | 0.497 | 0.482 | 0.488 | 0.008 | 1.670 |
| day 5 (pg/mL) | 0.472 | 0.462 | 0.464 | 0.466 | 0.005 | 1.136 |

repeatability test of the Lumipulse G600II using 3 aliquots of a patient diagnosed with Alzheimer’s Disease (Positive control, PC) tested in 5 different days. p-tau217, phosphorylated tau 217; SD, Standard Deviation; CV, Coefficient of Variation; 95%CI

**Supplementary Table 3. Repeatability test Level 1, L1 (5 replicates X 5 testing days)**

| p-tau217 LUMIPULSE | L1-1 | L1-2 | L1-3 | L1-4 | L1-5 | MEAN | SD | CV(%) |
| --- | --- | --- | --- | --- | --- | --- | --- | --- |
| day 1 (pg/mL) | 0.508 | 0.457 | 0.448 | 0.461 | 0.465 | 0.468 | 0.023 | 4.989 |
| day 2 (pg/mL) | 0.448 | 0.502 | 0.504 | 0.502 | 0.453 | 0.482 | 0.029 | 5.944 |
| day 3 (pg/mL) | 0.439 | 0.44 | 0.467 | 0.498 | 0.506 | 0.470 | 0.031 | 6.686 |
| day 4 (pg/mL) | 0.511 | 0.503 | 0.457 | 0.472 | 0.495 | 0.488 | 0.022 | 4.608 |
| day 5 (pg/mL) | 0.492 | 0.469 | 0.469 | 0.502 | 0.453 | 0.477 | 0.020 | 4.132 |

repeatability test of the Lumipulse G600II using 5 commercial QC available products (Level 1, L1) tested in 5 different days. p-tau217, phosphorylated tau 217; SD, Standard Deviation; CV, Coefficient of Variation; 95%CI

**Supplementary Table 4. Repeatability test Level 2, L2 (5 replicates X 5 testing days)**

| p-tau217 LUMIPULSE | L2-1 | L2-2 | L2-3 | L2-4 | L2-5 | MEAN | SD | CV(%) |
| --- | --- | --- | --- | --- | --- | --- | --- | --- |
| day 1 (pg/mL) | 3.922 | 3.637 | 3.668 | 3.955 | 3.670 | 3.770 | 0.154 | 4.096 |
| day 2 (pg/mL) | 3.675 | 3.583 | 3.840 | 3.738 | 3.845 | 3.736 | 0.112 | 2.987 |
| day 3 (pg/mL) | 3.643 | 3.9 | 3.696 | 3.956 | 3.984 | 3.836 | 0.156 | 4.065 |
| day 4 (pg/mL) | 3.942 | 3.702 | 3.864 | 4.001 | 3.789 | 3.860 | 0.119 | 3.083 |
| day 5 (pg/mL) | 3.815 | 3.75 | 3.66 | 3.959 | 3.726 | 3.782 | 0.113 | 2.999 |

repeatability test of the Lumipulse G600II using 5 commercial QC available products (Level 2, L2) tested in 5 different days. p-tau217, phosphorylated tau 217; SD, Standard Deviation; CV, Coefficient of Variation; 95%CI

**Supplementary Table 5. Precision results of the commercial QC, level 1 (L1) and level 2 (L2), and positive (PC) and negative controls (NC)**

| **Precision** | **L1** | **L2** | **NC** | **PC** |
| --- | --- | --- | --- | --- |
| **General mean** | 0.477 | 3.797 | 0.184 | 0.479 |
| **DS Laboratory** | 0.024 | 0.129 | 0.007 | 0.011 |
| **CV within Laboratory** | 5.080 | 3.387 | 3.749 | 2.340 |
| **CV between run** | 5.340 | 3.490 | 2.280 | 1.310 |

**Supplementary Table 6.** **Participants’ characteristics and plasma biomarkers assessed by Lumipulse and SIMOA platforms for MCI.**

|  | **AD**  **(N=112)** | **NDD**  **(N=45)** | **p-value** |
| --- | --- | --- | --- |
| **Age** | 72.698 (6.958) | 68.181 (7.456) | 0.001 |
| **Sex (F:M)** | 70:42 | 22:23 | 0.117 |
| **MMSE** | 24.612(3.571) | 28.500 (1.202) | <0.001 |
| **AD CSF core biomarkers** | | | |
| **t-tau (pg/mL)** | 679.783 (319.827) | 277.707 (304.062) | <0.001 |
| **p-tau181 (pg/mL)** | 116.227 (61.922) | 37.224 (13.079) | <0.001 |
| **Aβ42 (pg/mL)** | 473.747 (154.606) | 906.231 (303.042) | <0.001 |
| **Plasma biomarkers** | | | |
| **Plasma p-tau217 (L) (pg/mL)** | 0.751 (0.468) | 0.166 (0.103) | <0.001 |
| **Plasma p-tau217 (S) (pg/mL)** | 1.108 (0.554) | 0.351 (0.221) | <0.001 |

**Supplementary Table 7. Area Under the Curve (AUC), 95% Confidence Intervals (CI), Sensitivity, Specificity and Youden cut-off for ROC analysis on p-tau217, phosphorylated tau 217 tested on Lumipulse (L) and SIMOA (S) for AD-MCI and NDD-MCI.**

|  |  | **AUC** | **95%CI** | **Sensitivity** | **Specificity** | **Youden cut-off** |
| --- | --- | --- | --- | --- | --- | --- |
| **p-tau217 (L)** | **AD-MCI *vs* HC** | 0.960 | 0.936-0.985 | 0.918 | 0.933 | 0.251 |
|  | **AD *vs* NDD-MCI** | 0.946 | 0.911-0.981 | 0.882 | 0.848 | 0.287 |
| **p-tau217 (S)** | **AD-MCI *vs* HC** | 0.960 | 0.936-0.985 | 0.918 | 0.922 | 0.524 |
|  | **AD *vs* NDD-MCI** | 0.934 | 0.893-0.976 | 0.864 | 0.848 | 0.599 |
